# Independent Assessment of a Point of Care HCV RNA Test by Laboratory Analytical Testing and a Prospective Field Study in the U.S

**DOI:** 10.1101/2025.05.01.25326028

**Authors:** Lesley S. Miller, Anuradha Rao, Karlyn Tunnell, Yun F. Wang, Richard Parsons, Kaleb McLendon, Leda Bassit, Mahi Patel, Heather B. Bowers, Courtney Sabino, Thanuja Ramachandra, Farzan Saeed, Raymond F. Schinazi, Wilbur Lam, Julie A Sullivan, Shelly-Ann Fluker

## Abstract

Improvements in HCV testing, including Point of Care (POC) HCV RNA tests, are necessary to eliminate HCV. With this goal in mind, we established methods to collect capillary whole blood (CWB) via fingerstick, ensured its stability in a microtainer, and determined limit of detection (LoD) for various HCV genotypes. Next, we conducted a prospective study where CWB samples were collected from a cohort of 109 adult subjects at a safety-net hospital and tested for HCV RNA using the Xpert® HCV test on the GeneXpert® Xpress system and the cobas^®^ HCV platform. We consistently obtained 250 μl CWB which was stable for up to 5 hours in the microtainer. Laboratory LoD studies demonstrated that the Xpert® HCV test could detect most HCV genotypes to <100 IU/ml. In the prospective clinical study, 89 subjects (82%) with valid Xpert® and cobas^®^ HCV comparator results were analyzed. Using Xpert®, 16 out of 89 (18%) subjects had detectable HCV RNA and 73 (82%) had undetectable HCV RNA. Using cobas, 17 out of 89 (19%) participants had detectable HCV RNA, and 72 (81%) were undetectable. One sample was detectable on cobas but not Xpert®, yielding a sensitivity of 94% and specificity of 100%. This study demonstrates the feasibility of HCV RNA testing at POC using CWB obtained by fingerstick and provides preliminary data on the accuracy of the Xpert® HCV test performed by untrained operators in a CLIA-waived setting.

**Summary:** This is the first U.S. study to evaluate both the laboratory analytic performance as well as the clinical performance in a prospective field trial of the point of care Xpert® HCV test.

## Introduction

Hepatitis C virus (HCV) infection is a major public health concern in the United States. HCV is responsible for over 13,000 deaths and 70,000 new infections per year, with incidence increasing steadily since 2014.[1] Although novel therapies result in cure in >95% of those treated, HCV treatment rates remain suboptimal, with less than 35% of individuals with known infection receiving treatment, even among insured populations.[2] Deficiencies in screening, healthcare access, and affordability of medications continue to be the main drivers of treatment disparities.[3]

To address this public health crisis, a United States National Hepatitis C Elimination Program has been proposed. The program rests on 3 pillars, the first of which is accelerating the availability of point-of-care (POC) HCV tests.[4] Traditional standard of care (SOC) HCV testing requires two steps to be performed in a laboratory: detection of HCV antibodies followed by confirmation of active infection through HCV RNA testing. As this testing process is lengthy and treatment is not initiated until after RNA testing, many patients are lost to follow-up before RNA testing is completed, leading to a substantial gap in the care cascade.[5] In contrast, POC HCV RNA testing allows for rapid diagnosis of active infection in a single step from a fingerstick blood sample. POC HCV RNA testing, including testing of capillary whole blood (CWB) obtained by fingerstick has been successful in Europe, Australia, Tanzania and Switzerland, but had not yet been validated and approved in the United States.[6–9]

Patients have found POC testing to be an acceptable alternative to SOC testing, with over half preferring POC tests.[10,11] In addition, POC HCV RNA tests have a high diagnostic accuracy, with a sensitivity and specificity of 99% for HCV RNA detection and 100% for HCV RNA quantification when compared to SOC.[12] For these reasons, the World Health Organization (WHO) recommends POC testing as an alternate to SOC particularly in high risk populations.[13] This is largely due to the reduced time to initiation of treatment with POC tests, an average of 19 days from positive test to the beginning of treatment, compared to 64 days with SOC testing. POC testing also leads to increased treatment uptake, with 77-81% of individuals initiating treatment compared to 53% with SOC.[6]

As a foundational step in the launch of the National Hepatitis C Elimination Program, we conducted the first U.S. independent assessment of the POC Cepheid Xpert® HCV test. The Xpert® HCV test, performed using the Cepheid GeneXpert® Xpress system, is an automated qualitative *in vitro* reverse transcription polymerase chain reaction (RT-PCR) test for the detection of HCV RNA using CWB from a fingerstick sample. Our study is unique in that it included both laboratory analytical testing and a prospective field study. This is the first study to evaluate the stability of CWB under different conditions in a Microtainer that has not been used previously for CWB HCV testing but is readily available in the United States from Becton Dickinson. This study describes the first side-by-side Probit LoD study of Xpert® HCV test for its ability to detect all 6 of the most common HCV genotypes in venous whole blood (VWB) versus CWB. Based on FDA guidance, comparable detection in both types of matrices was identified as critical for Cepheid to receive de novo marketing authorization and CLIA waiver approval. Finally, the accuracy and feasibility of HCV RNA POC testing using the Xpert® HCV by untrained users in a U.S. CLIA-waived setting with high HCV prevalence was assessed for the first time.

## Methods

### Human Subjects

The protocol was approved by the Emory University Institutional Review Board No. 6542 and the Grady Research Oversight Committee. All research was conducted in accordance with both the Declarations of Helsinki and Istanbul, and written consent was given by all subjects.

### Laboratory Analytical Testing

#### CWB Collection

For this study, a minimum volume of 250μl CWB from fingerstick was required when using the microtainer from Becton Dickinson (BD catalog number 365974). We used a modified protocol from the CDC and followed the Instructions for Use (IFU) for Xpert® HCV test.[14,15] An Accu-Chek Safe-T-Pro-Plus (catalog number 03448622001) or blue BD lancet (catalog number 366594) was used. After collection, the microtainer was capped and blood mixed by inverting 10 times.

#### Stability of CWB in BD Microtainer

Venous Whole Blood (VWB) was obtained from six consented donors for HCV screening with the cobas^®^ HCV test for the 5800/6800/8800 Systems. All six donors were negative. Five HCV genotype 1b serum or plasma clinical samples were obtained from the Grady Hospital HCV biorepository. HCV RNA levels were determined using cobas^®^ HCV **(Supplementary Table 1)**. CWB from HCV negative donors were spiked with HCV genotype 1b positive samples at ∼3X LOD, after which they were stored in the microtainer for 0, 4 and 5 hours at 2°C and 30°C. Samples were analyzed using the Xpert® HCV test and results were recorded.

#### Analytical Limit of Detection (LOD) of HCV Genotypes 1-6 in CWB and VWB

HCV genotypes 1-6 were obtained from the sources listed in **Supplementary Table 2**. LOD range finding (replicates of 5) and LOD confirmation (replicates of 20) were carried out for each genotype. Donor distributions, Probit calculation, and other details are described in **Supplementary Methods, and in Supplementary Tables 3 and 4**.

Studies with VWB were conducted by MRIGlobal (Kansas City, MO), with details indicated in supplementary methods.

### Prospective Field Study

#### Study Population

We performed a prospective clinical study comparing HCV RNA POC testing using the Xpert® HCV test on the Xpress system with SOC HCV RNA testing using the cobas^®^ HCV test. The study was performed at the Grady Health System (GHS) and the Emory University Research Laboratory (Atlanta, GA, USA). Participants were enrolled from the GHS Primary Care Center and Grady Liver Clinic from October 2023 through January 2024. The Liver Clinic was selected for recruitment based on high prevalence of HCV exposure (7%) and chronic infection (3%).[16] Adults aged ≥ 18 years were eligible to participate, and signs/symptoms of HCV infection were not required for enrollment. We excluded persons who were unable to provide consent, pregnant, incarcerated and/or non-English-speaking. Chart reviews and subject interviews were performed to collect demographic and comorbidity data.

#### Procedures

Two staff members previously untrained on the Xpress system performed all study interventions using instructions provided by the manufacturer. Staff attempted to collect 250μl CWB via fingerstick and venous blood samples from each subject. The CWB in this study was collected in a BD microtainer, a K2EDTA coated collection device, not previously cleared for molecular diagnostic use. Venipuncture was performed prior to fingerstick, and CWB samples were not collected from subjects with unsuccessful venipuncture, as venous samples were required for final population inclusion and analysis. Immediately after collection, a pipette provided by the manufacturer was used to transfer 100 μl of CWB into the Xpert® HCV test cartridge for analysis on the Xpress system. Four samples could be analyzed concurrently on the system with a run time of approximately 1 hour. Results were reported as HCV detected, HCV not detected, Error or No Result. Venous blood samples were tested for HCV antibodies and HCV RNA using the cobas^®^ HCV platform in the Emory University laboratory and the Alinity m system platform in the GHS Clinical Microbiology and Molecular Diagnostics Laboratory. Venous samples were also tested for hepatitis B virus (HBV) surface antigen, surface antibody and core antibody using the Alinity i platform in the GHS Clinical Immunology Laboratory.

#### Statistical Analysis

All statistical analyses were performed using R v4.4.0,[17] and all counts and tables were produced using the dplyr and r2rtf packages.[18,19] 95% confidence intervals were calculated using the DescTools package.[20] See supplemental materials for additional details.

## Results

### Stability of CWB in the BD microtainer

CWB collected in the BD microtainer was spiked at 3x LoD with individual HCV genotype 1a clinical samples and one CWB sample was spiked with HCV negative VWB from a confirmed HCV negative donor. Samples were incubated at indicated temperatures and times. Results in **Table 1** demonstrate that CWB samples spiked with HCV negative VWB gave negative results at all times and temperatures. Samples spiked with genotype 1a gave positive results (HCV DETECTED) at all times and temperatures. HCV positive and HCV negative CWB in the BD microtainer was stable for up to 5 hours between 2°C and 30°C.

**Table 1.**
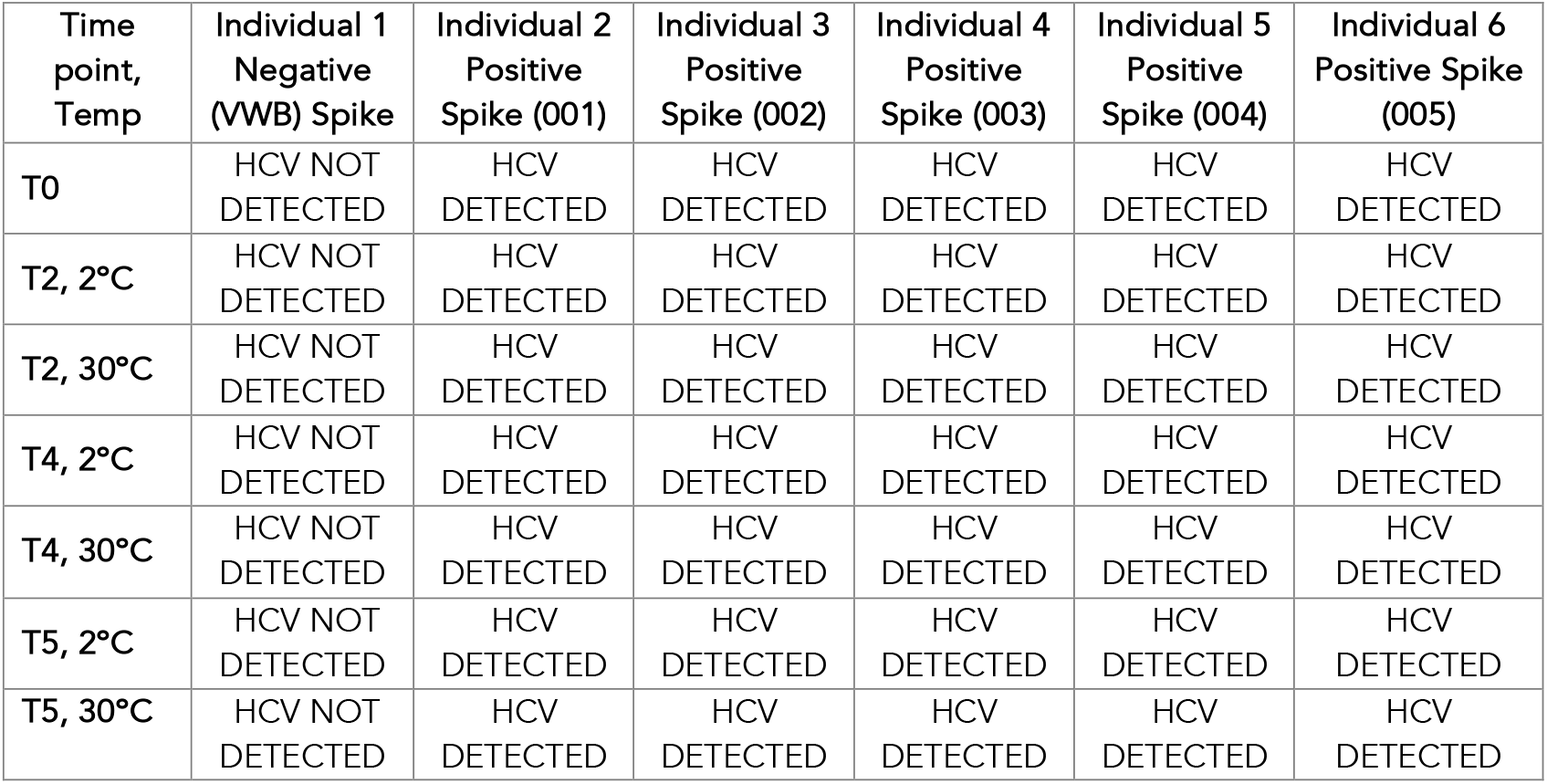
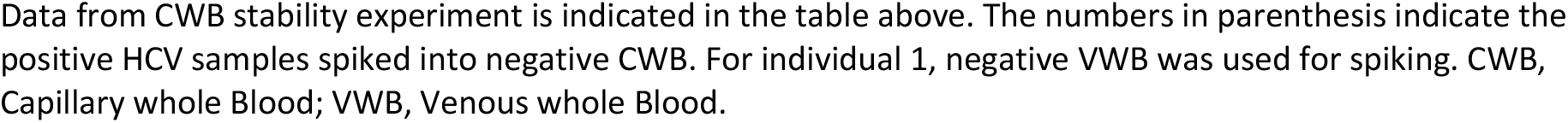
Xpert® HCV Capillary Whole Blood Specimen Stability Data.

| Time point, Temp | Individual 1 Negative (VWB) Spike | Individual 2 Positive Spike (001) | Individual 3 Positive Spike (002) | Individual 4 Positive Spike (003) | Individual 5 Positive Spike (004) | Individual 6 Positive Spike (005) |
| --- | --- | --- | --- | --- | --- | --- |
| T0 | HCV NOT DETECTED | HCV DETECTED | HCV DETECTED | HCV DETECTED | HCV DETECTED | HCV DETECTED |
| T2, 2°C | HCV NOT DETECTED | HCV DETECTED | HCV DETECTED | HCV DETECTED | HCV DETECTED | HCV DETECTED |
| T2, 30°C | HCV NOT DETECTED | HCV DETECTED | HCV DETECTED | HCV DETECTED | HCV DETECTED | HCV DETECTED |
| T4, 2°C | HCV NOT DETECTED | HCV DETECTED | HCV DETECTED | HCV DETECTED | HCV DETECTED | HCV DETECTED |
| T4, 30°C | HCV NOT DETECTED | HCV DETECTED | HCV DETECTED | HCV DETECTED | HCV DETECTED | HCV DETECTED |
| T5, 2°C | HCV NOT DETECTED | HCV DETECTED | HCV DETECTED | HCV DETECTED | HCV DETECTED | HCV DETECTED |
| T5, 30°C | HCV NOT DETECTED | HCV DETECTED | HCV DETECTED | HCV DETECTED | HCV DETECTED | HCV DETECTED |

Data from CWB stability experiment is indicated in the table above. The numbers in parenthesis indicate the positive HCV samples spiked into negative CWB. For individual 1, negative VWB was used for spiking. CWB, Capillary whole Blood; VWB, Venous whole Blood.

### Xpert® HCV Limit of detection

The LoD for Genotypes 1 to 6 and ranged between 35 IU/ml for genotype 1a to 136.4 IU/ml for genotype 5 in CWB. The range in VWB was from 43.8 for genotype 1b to 126.2 for genotype 6 (**Table 2**).

**Table 2.** Xpert® HCV LoD estimated by Probit. The number of datapoints used for each LoD calculation are indicated.

| HCV Genotype | Probit LoD in IU/ml (95% Confidence Interval),<br>Number of datapoints used for LoD calculation |  |
| --- | --- | --- |
|  | CWB LoD | VWB LoD |
| 1a | 35.0 (22.3 - 54.9), 90 | 74.2 (39.9 - 138.0), 85 |
| 1b | 41.5 (29.8 - 57.9), 95 | 43.8 (30.3 – 63.2), 110 |
| 2b | 89.2 (47.7 - 166.8), 100 | 66.7 (41.1 – 108.3), 120 |
| 3a | 58.2 (39.0 - 86.7), 125 | 75.3 (50.6 - 112), 125 |
| 4 | 32.2 (21.1 - 49.1), 120 | 39.5 (26.1 – 59.9), 80 |
| 5 | 136.4 (83.2 - 223.4), 145 | 119.1 (74.3 – 190.7), 100 |
| 6 | 83.8 (57.2 - 122.8), 120 | 126.2 (71.3 – 223.4), 120 |

Comparison of LoD obtained for all six HCV genotypes using CWB and VWB. Numbers in parenthesis indicate 95% confidence interval. The numbers of datapoints used in calculation of Probit LoD is indicated. LoD, Limit of Detection; HCV, Hepatitis C Virus; CWB Capillary Whole Blood; VWB, Venous whole Blood.

### Feasibility

The two untrained operators successfully incorporated POC HCV RNA testing on the Xpress system into clinical workflow. Of 109 subjects enrolled, 12 were excluded because of unsuccessful venipuncture (**Fig 1**). The remaining 97 subjects were eligible for CWB sample collection for Xpert® HCV testing. 89 CWB samples (92%) yielded valid results on the Xpress system. The remaining 8 samples (8%) had results that were “Error” or “No Result,” either due to insufficient CWB sample transferred to the cartridge (6 samples), clot formation in the collection device after CWB sample collection (1 sample), or user error (user attempted to run the sample before the control run was complete, 1 sample). These issues arose most commonly at the onset of the study, with 7 of the 8 invalid results occurring during the first week of sample collection. This was attributed to staff becoming accustomed to fingerstick sample collection and system operation.

**Figure 1.**
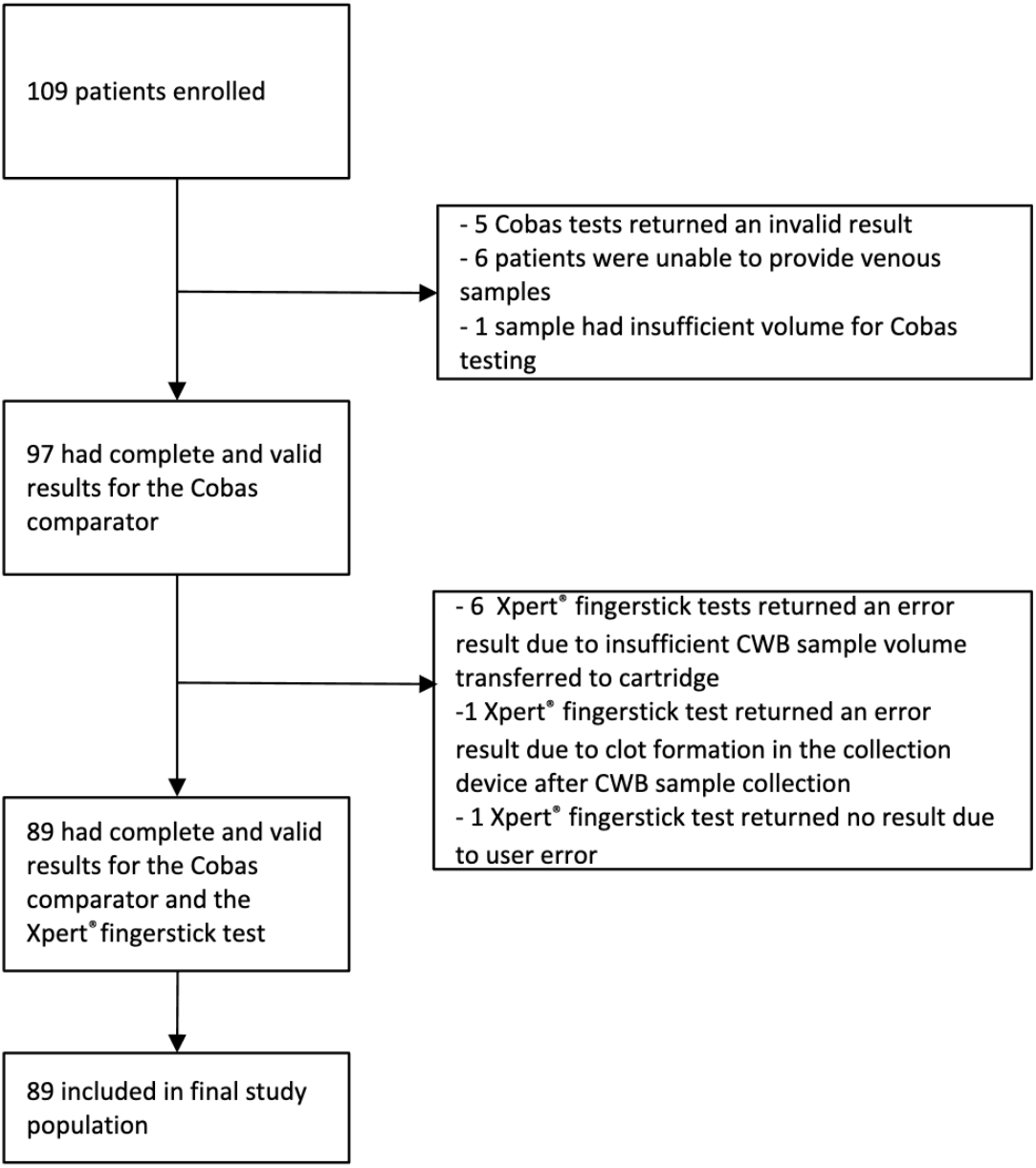
Study Flow Diagram. 109 subjects were initially recruited, of which 12 subjects were excluded because of invalid or incomplete samples on the Cobas comparator and 8 because of invalid or incomplete Xpert® test results. This left 89 subjects in the final study population. CWB, Capillary Whole Blood.

### Subject Characteristics

Of 109 subjects enrolled in the study, 89 had both valid Xpert® HCV and cobas^®^ HCV results and were included in the final study population for analysis (**Fig 1**). As shown in **Table 3**, most of the subjects were male and black/African American (65% and 82%, respectively). 72 subjects (81%) had been previously tested for HCV antibodies, 37 (51%) of these had positive results, and 19 of these 37 (51%) had detectable HCV RNA. Regarding HBV, 33 (37%) had positive HBV surface antibodies, one subject had a positive HBV surface antigen and 9 (10%) had isolated core antibody status. Less than 5% of the subjects had HIV infection. 44% had current or prior alcohol use, and 12% had current or prior injection drug use history.

**Table 3.**
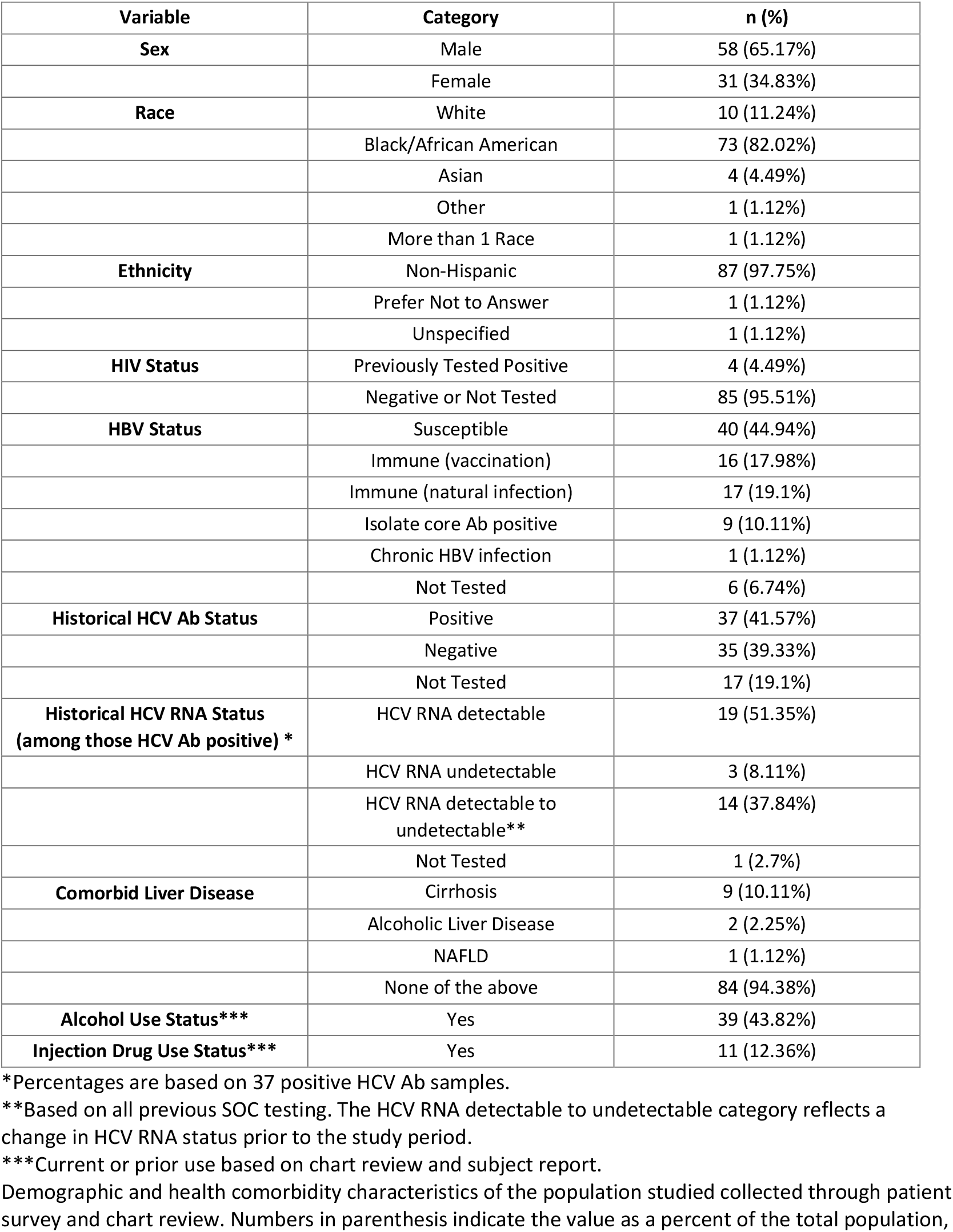
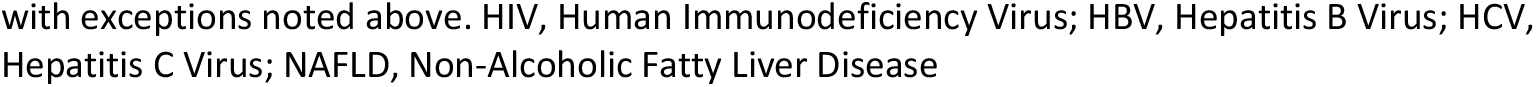
Characteristics of the Study Population.

| **Variable** | **Category** | **n (%)** |
| --- | --- | --- |
| **Sex** | Male | 58 (65.17%) |
|  | Female | 31 (34.83%) |
| **Race** | White | 10 (11.24%) |
|  | Black/African American | 73 (82.02%) |
|  | Asian | 4 (4.49%) |
|  | Other | 1 (1.12%) |
|  | More than 1 Race | 1 (1.12%) |
| **Ethnicity** | Non-Hispanic | 87 (97.75%) |
|  | Prefer Not to Answer | 1 (1.12%) |
|  | Unspecified | 1 (1.12%) |
| **HIV Status** | Previously Tested Positive | 4 (4.49%) |
|  | Negative or Not Tested | 85 (95.51%) |
| **HBV Status** | Susceptible | 40 (44.94%) |
|  | Immune (vaccination) | 16 (17.98%) |
|  | Immune (natural infection) | 17 (19.1%) |
|  | Isolate core Ab positive | 9 (10.11%) |
|  | Chronic HBV infection | 1 (1.12%) |
|  | Not Tested | 6 (6.74%) |
| **Historical HCV Ab Status** | Positive | 37 (41.57%) |
|  | Negative | 35 (39.33%) |
|  | Not Tested | 17 (19.1%) |
| **Historical HCV RNA Status**<br>**(among those HCV Ab positive) \*** | HCV RNA detectable | 19 (51.35%) |
|  | HCV RNA undetectable | 3 (8.11%) |
|  | HCV RNA detectable to undetectable** | 14 (37.84%) |
|  | Not Tested | 1 (2.7%) |
| **Comorbid Liver Disease** | Cirrhosis | 9 (10.11%) |
|  | Alcoholic Liver Disease | 2 (2.25%) |
|  | NAFLD | 1 (1.12%) |
|  | None of the above | 84 (94.38%) |
| **Alcohol Use Status\*\*\*** | Yes | 39 (43.82%) |
| **Injection Drug Use Status\*\*\*** | Yes | 11 (12.36%) |

\*Percentages are based on 37 positive HCV Ab samples.
\*\*Based on all previous SOC testing. The HCV RNA detectable to undetectable category reflects a change in HCV RNA status prior to the study period.
\*\*\*Current or prior use based on chart review and subject report.
Demographic and health comorbidity characteristics of the population studied collected through patient survey and chart review. Numbers in parenthesis indicate the value as a percent of the total population,
with exceptions noted above. HIV, Human Immunodeficiency Virus; HBV, Hepatitis B Virus; HCV, Hepatitis C Virus; NAFLD, Non-Alcoholic Fatty Liver Disease

**Table 4.** POC Xpert® HCV Test Performance Compared to cobas^®^ HCV Venous Blood Testing.

|  |  | Cobas® HCV Venous Blood Test Result |  |  |
| --- | --- | --- | --- | --- |
|  |  | HCV RNA Detected | HCV RNA Not Detected | Total |
| Xpert® HCV Test Result | HCV RNA Detected | 16 | 0 | 16 |
|  | HCV RNA Not Detected | 1 | 72 | 73 |
|  | Total | 17 | 72 | 89 |

Data in the above table indicate performance of the Xpert® HCV test with sensitivity of 94% (95% CI 71.3%, 99.9) and Specificity of 100% (95% CI 95.0%, 100%). CI, Confidence Interval.

### Xpert® HCV Test Performance

Using the Xpert® HCV test, 16 of 89 (18%) subjects had an HCV RNA detected result, while the remaining 73 (82%) had an HCV RNA not detected result. The results were almost identical on the comparator cobas^®^ HCV assay: 17 out of 89 (19%) had HCV RNA detected, and the remaining 72 (81%) had HCV RNA not detected. Only one sample was detectable on the cobas^®^ HCV assay but not on the Xpert® HCV test, giving a sensitivity of 94% (95% CI 71.3%, 99.9) and specificity of 100% (95% CI 95.0%, 100%) **(Table** 4). This subject had an HCV RNA level of 18.7 IU/mL on the cobas^®^ HCV assay and <12 IU/mL on the Alinity m assay.

## Discussion

Our study is the first in the U.S. to report both the laboratory analytical performance (sample stability and LoD) as well as the preliminary clinical performance of the POC Xpert® HCV test using the GeneXpert® Xpress system. In addition, we were able to assess a novel workflow that used CWB collection for POC RNA testing. This study provided insight into the Xpert® HCV test’s utility for diagnosing active HCV infection and ultimately laid the groundwork for the clinical trial that informed FDA approval of the test.

Our measured specificity of 100% is consistent with prior studies performed outside of the U.S. However, our sensitivity of 94% was slightly lower than the sensitivity of 99% reported in these studies.[12,21] Our study differed from these studies in several aspects. First, ours was an assessment of a qualitative HCV RNA test under development for HCV diagnostic purposes only, in contrast to a quantitative test for on-treatment monitoring or test of cure after treatment. Second, a key component of this study was testing a novel workflow using a microtainer for specimen collection and a pipette provided with the Xpert® HCV test for specimen transfer. Finally, our study differs in setting and patient population.

Our study had a 94% sensitivity due to one false-negative sample. Notably, this sample had an extremely low viral load of 18.6 IU/mL when measured using SOC assays. Given that the Xpert® HCV test has a higher LoD of 35 to 136.4 IU/mL, this result was not surprising. Prior analyses examining the optimal LoD for a POC HCV test have determined that detecting viral loads greater than 1,300 IU/mL would be acceptable. This limit would detect 97% of those with chronic infection while still allowing for an affordable and portable test. Further lowering the LoD to 214 IU/mL would detect approximately 99% of those with chronic infection.[22] The Xpert® HCV test exceeds this value, providing reliable detection of clinically significant HCV viremia.[23] In our study, this subject’s HCV RNA was not detected at Quest laboratories and was below the lower limit of quantification on the Alinity system, tests which were performed for clinical care.

In our study, the Xpert® HCV test produced an error or invalid result in 8.2% of samples, which is consistent with other studies examining the test.[12] We did observe a learning curve regarding the operation of the device and collection of fingerstick samples and anticipate that failures due to inadequate CWB volume would decrease with user experience. For example, test operators improved their CWB collection technique after the first week of the study by warming the subjects’ hands prior to sample collection (using repetitive motion, warm water or warming gel packs), utilizing the maximum depth on the lancet depth setting, and ensuring complete sample transfer from pipette to cartridge by inserting the pipette to the maximum depth of the cartridge. After practice, all operators were consistently able to collect at least 250 μL of CWB from volunteers for laboratory LoD studies and from the subject population described here. These techniques were important in informing sample collection recommendations for the subsequent clinical trial for the Xpert® HCV test.

The reliability of the Xpert® HCV test is particularly notable given the patient population tested in our study. We enrolled patients from an HCV treatment clinic, which explains the high percentage of subjects (19.1%) with active HCV infection compared to an estimated 1% of individuals in the general population.[1] An additional 19.1% of our subjects had prior HCV infection that was treated or cleared. The reliable performance of the Xpert® HCV test in individuals with past HCV infection but undetectable HCV RNA underscores the utility of this test. Our study participants also reflected the medical complexity of those most at risk for HCV infection, with many having concurrent HIV infection, prior HBV infection, and/or a diagnosis of liver disease.[24,25] Notably, 30.3% of our participants had either active or prior HBV infection. Studies evaluating the performance of POC tests in the presence of HBV or HIV co-infection are limited, and we observed accurate test performance regardless of co-infection status.[21] Finally, we noted a high venipuncture failure rate in this population with a high prevalence of active or prior HCV infection. Challenging venous access and inability to acquire samples for testing is a major barrier to HCV diagnosis, especially among people who inject drugs, a key group to target for testing to meet HCV elimination goals. This underscores the need for a POC HCV RNA test available for use in individuals with difficult venous access.

This study had some limitations. Our sample size was small, with 109 subjects enrolled and 89 ultimately included in the analysis. In addition, the confidence intervals for the test sensitivity were wide. The study included a small number of participants with a history of intravenous drug use (IVDU), with only 12.4% reporting current or prior IVDU. This group is particularly at risk for HCV infection, and the prevalence of IVDU associated with the opioid epidemic has driven the increase in HCV infections within the past 10 years.[26] However, we do not have evidence to believe that the Xpert® HCV test would perform differently in a population with higher IVDU prevalence. ⍰

Despite these limitations, the Xpert® HCV test has great potential to both expand and enhance the settings in which HCV screening can be implemented. This study shows that it is possible to hold a CWB sample in the BD microtainer for up to 5 hours prior to starting the Xpert® HCV test, which offers additional flexibility beneficial in carceral, substance use disorder treatment, and mobile testing programs settings and community health screening events, where there may be physical separation between the site of sample collection and analysis or a delay in testing. Stability at the relatively high temperature of 30°C can be useful where refrigeration is not readily accessible or where there is frequent loss of electricity. The ability to collect at least 250 μL CWB, and its stability for up to 5 hours will enable a potential for a second test if a run is interrupted due to a power failure or if the first test result is invalid.

This was the first extensive laboratory study involving several hundred datapoints conducted using both CWB and VWB showing no differences in detection of a target in a nucleic acid-based detection assay when using either as the matrix. Demonstrating matrix equivalency was an important consideration based on FDA guidance for approval to receive de novo marketing authorization and CLIA waiver from the FDA. In addition to detection of HCV, the techniques and methods described here for laboratory HCV CWB LoD determination will be useful for manufacturers to develop tests for detection of other blood borne pathogens.

Overall, this study provides valuable insights into the accuracy and clinical utility of the POC Xpert® HCV test performed on the GeneXpert® Xpress system and its integral role in HCV elimination in the United States. We found the device to be a reliable tool for the detection of HCV, with the benefit of allowing for accurate diagnosis in a fraction of the time compared to SOC. While more research is needed regarding incorporation in various clinical settings and evaluation in patients with a history of IVDU, POC HCV RNA testing is a promising tool to accelerate progress towards HCV elimination.

## Supporting information

Independent Assessment of a POC HCV RNA Test 4_17_25 Supplementary Information medRxiv.docx

## Data Availability

All data produced in the present work are contained in the manuscript

## List of Abbreviations

HCV: (hepatitis C virus)
SOC: (standard of care)
POC: (Point-of-care)
RT-PCR: (reverse transcription polymerase chain reaction)
GHS: (Grady Health System)
CWB: (capillary whole blood)
LoD: (limit of detection)
HBV: (hepatitis B virus)
IVDU: (intravenous drug use)

## Acknowledgments

We thank Eric Lai, Pamela Miller, Nira Pollock, Gail Radcliff, Emily Kennedy, Carlos Aparicio for their insightful comments and discussions during planning and execution of the study. We thank Evelyn Morales and Zianya Solis for their assistance in conducting the CWB studies and the Children’s Clinical and Translational Discovery Core, Department of Pediatrics, Emory University for phlebotomy to obtain VWB. We thank the MRIGlobal team (Tiffany Edwards, Hayden Smith, Kelsey Badger, Aidan Wilson, Nathan Garza and Karen Peltier) for their contributions to this study.

Presentation: Two abstracts describing the results presented here were presented at the 2024 American Association for the Study of Liver Disease (AASLD) Annual Meeting (The Liver Meeting), San Diego, CA.

## Author contributions

**Conceptualization:** LSM, AR, SAF, YFW, RP, KM, JAS. **Data Curation:** LSM, AR, SAF, RP, KM. **Formal Analysis:** LSM, AR, SAF, KT, YFW, KM, RP **Funding Acquistion:** RFS, WL. **Investigation:** LSM, AR, SAF, KM, LB, MP, HBB, CS, TR, FS. **Methodology:** LSM, AR, KLM, LB, JAS, SAF. **Project Administration:** LSM, AR, SAF. **Resources:** LSM, LB, SAF. **Supervision:** LSM, AR **Validation:** AR, MP, HBB, CS. **Writing-Original Draft Preparation:** LSM, AR, KT, SAF. **Writing-Review & Editing:** LSM, AR, KT, SAF

## Potential conflicts of interest

L.M. reports grant funding from Gilead Sciences and consulting fees from AbbVie. S.F. reports consulting fees from Roche. The other authors declare that they have no conflicts of interest.

Data not publicly available

Manuscript preparation: Cepheid provided assistance with study design and preparation of the manuscript.

