## Supplementary material for "Independent Assessment of a Point of Care HCV RNA Test by Laboratory Analytical Testing and a Prospective Field Study in the U.S": Independent Assessment of a POC HCV RNA Test 4_17_25 Supplementary Information medRxiv.docx

**Supplementary Table 1. Quantitation of HCV Genotype 1b clinical samples used for CWB spiking in sample stability study.**

| Patient ID (Anonymized) | Date Collected | Matrix | Cobas^®^ HCV Quantitation (IU/ml) |
| --- | --- | --- | --- |
| 001 | 10/2023 | Serum | 530,200 |
| 002 | 11/2023 | Plasma | 1,892,000 |
| 003 | 12/2023 | Plasma | 2,842,000 |
| 004 | 12/2023 | Plasma | 66,980 |
| 005 | 11/2023 | Plasma | 389,800 |

All samples used for data in Table 1 were quantified using Cobas 6800. The IU/ml values obtained from cobas 6800 were used to calculate the amount of material to spike into CWB -microtainer stability studies. IU/ml, International Units per milliliter; CWB, Capillary whole Blood.

**Supplementary Table 2. Quantitation, and source for HCV Genotypes used in LOD studies**

| Genotype | Sample Type | Stock Concentration (IU/ml) | Catalog Number | Source |
| --- | --- | --- | --- | --- |
| Genotype 1a | Human source material in lyophilized plasma, reconstitute in water | 233636 IU/ml (COA) | 18/184 | NIBSC (https://nibsc.org) |
| Genotype 1b | HCV positive serum | 3,908,000 IU/ml (as determined by cobas^®^ HCV | DLS16-72210 | Discovery LifeSciences (https://dls.com) |
| Genotype 2b | Serum SST | 1074446 IU/ml as determined by cobas^®^ HCV | 177102-13 | Biocollections Worldwide (https://www.biocollections.com) |
| Genotype 3a | Plasma | 11,180,000 IU/ml as determined by by cobas^®^ HCV | 141199-66 | Biocollections Worldwide |
| Genotype 4 | Plasma | 629,400 IU/ml as determined cobas^®^ HCV | 184895-13 | Biocollections Worldwide |
| Genotype 5 (used at Emory) | Plasma | 634400 IU/ml as determined by cobas^®^ HCV | 0HC0010100240009068 | FIND |
| Genotype 5 (used at MRI) | Plasma | 449000 IU/ml as determined by cobas^®^ HCV | OHC0010100240009071 | FIND |
| Genotype 6 | Plasma | 495,800 IU/ml as determined cobas^®^ HCV | DLS16-67657 | Discovery Life Science |

HCV genotypes used for this study were purchased from commercial sources. The concentration of genotype 1a was denoted in the COA. All other concentrations were determined using the cobas 6800. COA, Certificate of Analysis.

**Analytical Limit of Detection (LOD) of HCV Genotypes 1-6 in CWB and VWB.**

All HCV genotypes were quantitated using the cobas^®^ HCV test to verify starting concentrations used for each experiment, and these values were used for all spiking experiments. All donors who contributed CWB for LOD studies were first screened for HCV by cobas^®^ HCV and only HCV negative donors contributed. All stocks are diluted to 10X of original concentration using HCV negative VWB as matrix. 10X stocks are used to spike CWB for 1X HCV. The amount of VWB did not exceed 10% in each test sample. For all genotypes, an initial range finding study was carried out where 3 to 5 dilutions per genotype and 5 replicates per dilution were tested. Each replicate used CWB obtained from a different individual collected in a BD microtainer. An example is shown in **Supplementary Table 3**. Using a precision pipette, 108μl CWB was transferred to a microtainer tube, to which 12μl 10X HCV stock was added for a final concentration of 1X. After mixing by gentle pipetting, 100μl was transferred to the Xpert® HCV test cartridge and tested. Range finding was conducted for all genotypes prior to the LoD confirmation studies.

**Supplementary Table 3. Schematic followed for HCV range finding studies.**

| Range Finding (5 replicates) | | | | | |
| --- | --- | --- | --- | --- | --- |
| 100 IU/ml | Person 1 | Person 2 | Person 3 | Person 4 | Person 5 |
| 70 IU/ml | Person 1 | Person 2 | Person 3 | Person 4 | Person 5 |
| 45 IU/ml | Person 1 | Person 2 | Person 3 | Person 4 | Person 5 |
| 37.5 IU/ml | Person 1 | Person 2 | Person 3 | Person 4 | Person 5 |
| 18.75 IU/ml | Person 1 | Person 2 | Person 3 | Person 4 | Person 5 |
| 0 IU/ml | Person 1 | Person 2 | Person 3 | Person 4 | Person 5 |

Five different CWB samples were used for each replicate of every dilution. All concentrations were tested in replicates of five. CWB, capillary whole blood.

For LOD confirmation studies, 20 replicates were tested for each dilution. The dilutions were selected based on range finding data, and the lowest dilution that resulted in 5/5 positive was used as the starting concentration. For the 20 replicates, CWB from 5 donors were used such that no more than 4 replicates per dilution were from the same donor. The schematic shown in **Supplementary Table 4** was followed. Donor numbers, replicates distribution, and dilutions were recommended by the FDA.

**Supplementary table 4: Schematic followed for HCV range finding studies.**

| Initial conc. (IU/ml) | Final conc. (IU/ml) | LOD Confirmation (20 replicates) | | | | | | | | | | | | | | | | | | | |
| --- | --- | --- | --- | --- | --- | --- | --- | --- | --- | --- | --- | --- | --- | --- | --- | --- | --- | --- | --- | --- | --- |
| 1000 | **100** | P1 | P1 | P1 | P1 | P2 | P2 | P2 | P2 | P3 | P3 | P3 | P3 | P4 | P4 | P4 | P4 | P5 | P5 | P5 | P5 |
| 500 | **50** | P1 | P1 | P1 | P1 | P2 | P2 | P2 | P2 | P3 | P3 | P3 | P3 | P4 | P4 | P4 | P4 | P5 | P5 | P5 | P5 |
| 37.5 | **37.5** | P6 | P6 | P6 | P6 | P7 | P7 | P7 | P7 | P8 | P8 | P8 | P8 | P9 | P9 | P9 | P9 | P10 | P10 | P10 | P10 |
| 18.75 | **18.75** | P6 | P6 | P6 | P6 | P7 | P7 | P7 | P7 | P8 | P8 | P8 | P8 | P9 | P9 | P9 | P9 | P10 | P10 | P10 | P10 |

Five different CWB samples were used for every four replicates of every dilution. CWB, capillary whole blood.

Example of the scheme followed all LOD confirmation experiments. Persons (P) 1-5 and 6-10 will donate CWB. We will use 450 μl CWB+50 μl stock to make test samples. This will be enough for 4 replicates and overage for a pipetting margin. We used a minimum of 5 donors per LOD confirmation.

For LoD confirmation, we aimed for one dilution to generate a positive hit rate of 0.60 to 0.90 and one dilution to generate 100% positive results. If one of the initial 3 dilutions did not generate a 0.95 positive hit rate, a fourth dilution was tested. A minimum of three dilutions is necessary for conducting Probit analysis. Analyse-it® for Microsoft® Excel (v 6.15.4) Method Validation edition was used for calculating Probit LoD (positive rate at 0.95 probability, data points aggregated, and output log transformed per developer Statistical Reference Guide). Both range finding and LoD confirmation data were used for Probit analysis. All data generated irrespective of positive percent rate were included in Probit analysis. Data points entered into Probit calculations were identified by their concentrations (IU/mL) and assigned a nominal, binary indicator of either ‘0’ for a negative result or ‘1’ for a positive result. Invalid tests were assigned no indicator and were excluded from analysis.

Studies with VWB were conducted by MRIGlobal (Kansas City, MO). K2 VWB purchased from BioIVT and confirmed negative for HCV by Cobas HCV 5800/6800/8800 Systems was used as matrix. HCV genotypes indicated in Supplementary Table 2 were used to contrive VWB samples tested. All HCV genotypes used for CWB and VWB LoD studies were identical except Genotype 5 as the Foundation for Innovative New Diagnostics (FIND) does not allow transfer of samples between institutions.

**Supplementary Statistical Analysis**

Regarding historical HCV RNA results, the percentage of subjects with positive HCV RNA was calculated using a denominator of 37, which represented all subjects with a positive HCV antibody. For all other percentages, a denominator of 89 was used to reflect the number of participants in the final study population. To assess the performance of the Xpert® HCV test compared to cobas^®^ HCV RNA testing, a 2 × 2 table was constructed, and sensitivity and specificity were calculated and rounded to the nearest whole percentage. For all analyses, any study participant who was either missing a result or had an invalid/error/no result for either the Xpert® HCV test or the cobas^®^ HCV test was excluded.
